# Access to Episodic Primary Care: A Cross-sectional Comparison of Walk-in Clinics and Urgent Primary Care Centers in British Columbia

**DOI:** 10.1101/2022.05.25.22275602

**Authors:** Mary A. McCracken, Ian R. Cooper, Michee-Ana Hamilton, Jan Klimas, Cameron Lindsay, Sarah Fletcher, Morgan Price, Lindsay Hedden, Rita K. McCracken

**Affiliations:** Innovation Support Unit, Faculty of Medicine, University of British Columbia, Vancouver, BC, Canada; Department of Family Practice, University of British Columbia, Vancouver, BC, Canada; Faculty of Health Sciences, Simon Fraser University, Vancouver, BC, Canada

**Keywords:** Health services accessibility, ambulatory care facilities, after-hours care, primary health care, delivery of healthcare, health care quality, access, evaluation

## Abstract

**Background:** Walk-in clinics are non-hospital based primary care facilities that are designed to operate without appointments and provide increased healthcare access with extended hours. Urgent Primary Care Centres (UPCCs) were introduced to British Columbia (BC) in 2018 as an additional primary care resource that provides urgent, but not emergent care during extended hours. This study identifies publicly-reported access characteristics for episodic primary care in BC, and provides a clinic-level comparison between walk-in clinics and UPCCs.

**Methodology:** This cross-sectional study used publicly available data from all walk-in clinics and UPCCs in BC. A structured data collection form was used to record access characteristics from clinic websites, including business hours, weekend availability, attachment to a longitudinal family practice, and provision of virtual services.

**Results:** In total, 268 clinics were included in the analysis (243 walk-in clinics, 25 UPCCs). Of those, 225 walk-in clinics (92.6%) and two UPCCs (8.0%) were attached to a longitudinal family practice. Only 153 (63%) walk-in clinics offered weekend services, compared to 24 (96%) of UPCCs. Walk-in clinics offer the majority (8,968.6/ 78.4%) of their service hours between 08:00 and 17:00, Monday to Friday. UPCCs offer the majority (889.3/ 53.7%) of their service hours after 17:00.

**Conclusion:** Most walk-in clinics are associated with a longitudinal family practice and provide the majority of clinic services during typical business hours. More research that includes patient characteristics and care outcomes, analyzed at the clinic level, may be useful to support the optimization of episodic primary health care delivery.

## BACKGROUND

Walk-in clinics in Canada offer community-based, episodic care for patients with minor illnesses and injuries.^1–3^ They are typically non-hospital based facilities that operate without appointments, with some also providing after-hours care. Walk-in clinics have been operating in British Columbia (BC) since 1986^4^ and, at that time, were marketed as offering a convenient alternative to the emergency department for after-hours care.^5^ In BC, walk-in clinics are typically funded via fee for service physician remuneration^6^ and have the same College of Physicians and Surgeons practice standard as longitudinal primary care clinics.^7^ In 2018, urgent and primary care centers (UPCCs) were introduced to British Columbia, as a primary care resource designed to provide urgent, but not emergent, primary care services during extended hours.^8^ Unlike other walk-in clinics, these clinics receive funding via their regional health authority, offer allied health care services and physicians are paid via an alternative payment plan.^6^

Access to longitudinal primary care services after-hours has been shown to have variable impact on the volume of emergency department visits.^9–12^ However the impact of episodic primary care, like that provided by walk-in clinic and UPCCs, on emergency department usage is even less well understood. Some studies have found that urgent care facilities may either increase costs^13^ or have variable effects on emergency department crowding.^14^ Walk-in clinics have also been criticized for disrupting continuity of care which has been shown to decrease mortality, hospitalizations and healthcare costs.^2,15–17^ Since 2000, there have been reports of a family doctor shortage in BC^18^ and more widely in Canada.^19,20^ Approximately 15% of people, over the age of 12, living in BC are not attached to longitudinal primary care.^21^ In Canada, patients who are not attached to a longitudinal family practice appear to use walk-in clinics more frequently.^22^ It is possible resources used to provide episodic care have decreased resources available to provide longitudinal care.^23^

There are few published studies from the last decade that describe clinic numbers, characteristics or effects of episodic primary care in Canada.^24^ Such baseline description is important to be able to monitor changes over time and to understand how episodic primary care supports or detracts from longitudinal care. The objectives of this study are to describe and compare publicly-reported access characteristics including: clinic service hours, after hours availability, association with other primary care services, booking methods and appointment types. We will also describe the regional variation in number of clinic hours available per week, per 100,000 population in British Columbia.

## METHODS

### Study Design and Setting

This is a cross-sectional study of outpatient clinics providing episodic primary care in British Columbia (BC), Canada. BC has a single payer health care insurance program, and there are no direct to patient charges for primary care services delivered at walk-in clinics or UPCC’s.^25^ Physician payment is largely by fee for service, and there are only a few clinics where physicians are paid via a capitated model,^6^ that includes penalties if their patients go to walk-in clinics.^26^

### Data Collection and Sources

We used publicly available lists of clinic information including the BC Ministry of Health’s Walk-in Clinic List (Walk-in Clinic List) version: April 2021, ^27^ and the Urgent and Primary Care Clinic (UPCC) List, version: November 2021.^28^ The Walk-in Clinic List is maintained by staff at the Ministry of Health, reviewed for completeness by a data steward at the BC Data Catalogue.^29^ The listing includes: name of clinic, associated website, phone number, street address, as well as, additional data. The UPCC list^28^ is regularly updated on HealthLink BC which is the public-facing source of non-emergency health information and advice maintained by the BC Ministry of Health. This site provides a list of the names of UPCCs, organized by health authority, and a link to a clinic-specific website.^28^

The two lists were reviewed for duplication. If the Walk-in Clinic List or the UPCC listing did not include a website address, the clinic and street address were searched via Google. For any clinics with no self-managed website found in a google search, information would be extracted from one of the public, aggregate walk-in clinic websites, searched in order: Medimap^30^, SkiptheWaitingRoom^31^, Pathways^32^, and Cortico^33^.

A structured data collection form was used to review each clinic website for the variables of interest. Data were collected from walk-in clinic websites between May and June 2021 and from UPCC websites in both June and November 2021, by author MM. UPCC data was collected twice as 13 new UPCCs opened between the start and end of the study period.

Population data by health authority was retrieved from the BC government population estimates and projections website.^34^ Oversight of access to and quality of health care services in BC is delivered by 5 different geographically defined health authorities (there is also a First Nations Health Authority [FNHA] which is responsible for managing health programs and services for First Nations people in the entire province).^35^ The number of clinic service hours at walk-in clinic’s and UPCCs per 100,000 people are reported at this level because health system monitoring and funding is directed to health authorities.^36^

### Variables of Interest

#### Clinic service hours

Regular business hours were defined as 08:00-17:00, Monday to Friday. After-hours access was defined as before 08:00 and/or after 17:00 and/or after 20:00 Monday to Friday or any patient services available on weekends (Saturday and/or Sunday).

#### Association with other primary care services

A walk-in clinic was determined to be associated with a longitudinal family practice if the walk-in clinic website stated that they also provided attachment for longitudinal care or specifically described an association with a separate longitudinal clinic. This information was collected to help understand the possible relationship between longitudinal and episodic primary care provision. Presence of a co-located pharmacy was recorded as a proxy measure for presence of primary care services in addition to family doctor consultation, and that might be expected to be present for both walk-in clinic’s and UPCCs. This was determined if an adjacent pharmacy was listed on their website or observed by author MM through visualization on Google Maps.^37^

#### Appointment booking options and availability of virtual care options

Appointment booking information was collected from the clinic website. Ability to “walk-in” was determined if the website said the patient did not need to be previously known or did not require a booked appointment to attend the clinic for care. Types of “walk-in appointments” included walk-in only (where no other type of booking is available), phone booking, online and email bookings. Availability of virtual services was determined to exist if the clinic website listed options of telephone or video appointments. A change in access to services due to COVID was determined by the clinic’s website mentioning that walk-in services were no longer being provided or requesting patients to call in before coming to the clinic.

### Data Analysis

We used a Chi-square test to compare categorical variables, stratified by health authority and clinic type (walk-in clinics *vs*. UPCCs), and calculated descriptive statistics using the IBM Statistical Package for the Social Sciences software (V27.0; IBM). We accessed publicly available information and per article 2.2 of the Tri-Council Policy Statement 2 (2018) did not require review by an ethics board.^38^

## RESULTS

We included 268 clinics in our analysis, including: 243 walk-in clinics and 25 UPCCs. The Walk-in Clinic List (April 2021) listed 256 clinics, however, 13 were excluded, 12 because they were permanently closed and one did not list any service hours. Eighty of the walk-in clinics did not have a website listed, 42 walk-in clinic websites were found via a Google search and for the remaining 38 walk-in clinics data was collected from one of the aggregate websites:^30^ SkiptheWaitingRoom^31^, Pathways^32^, and Cortico^33^.

### Clinic service hours

There are 13,094 hours of episodic primary care available in a typical week in British Columbia (Table 1). Fourteen (5.8%) walk-in clinics provide patient services before 08:00, none of the UPCCs do. Sixteen (6.6%) walk-in clinics offer services after 17:00, compared to 22 (88%) of UPCCs, and 153 (63%) walk-in clinics are open on weekends, compared to 24 (96%) UPCCs. Walk-in clinics offered the majority (8,968.6/ 78.4%) of their service hours between 08:00 and 17:00, Monday to Friday. UPCCs offered the majority (889.3/ 53.7%) of their service hours after 17:00. Only 145 (59.7%) walk-in clinics, and none of the UPCCs listed the names and/or number of doctors who worked there (median 5 [Interquartile range (IQR) = 3.7]) (Table 1).

**Table 1:**
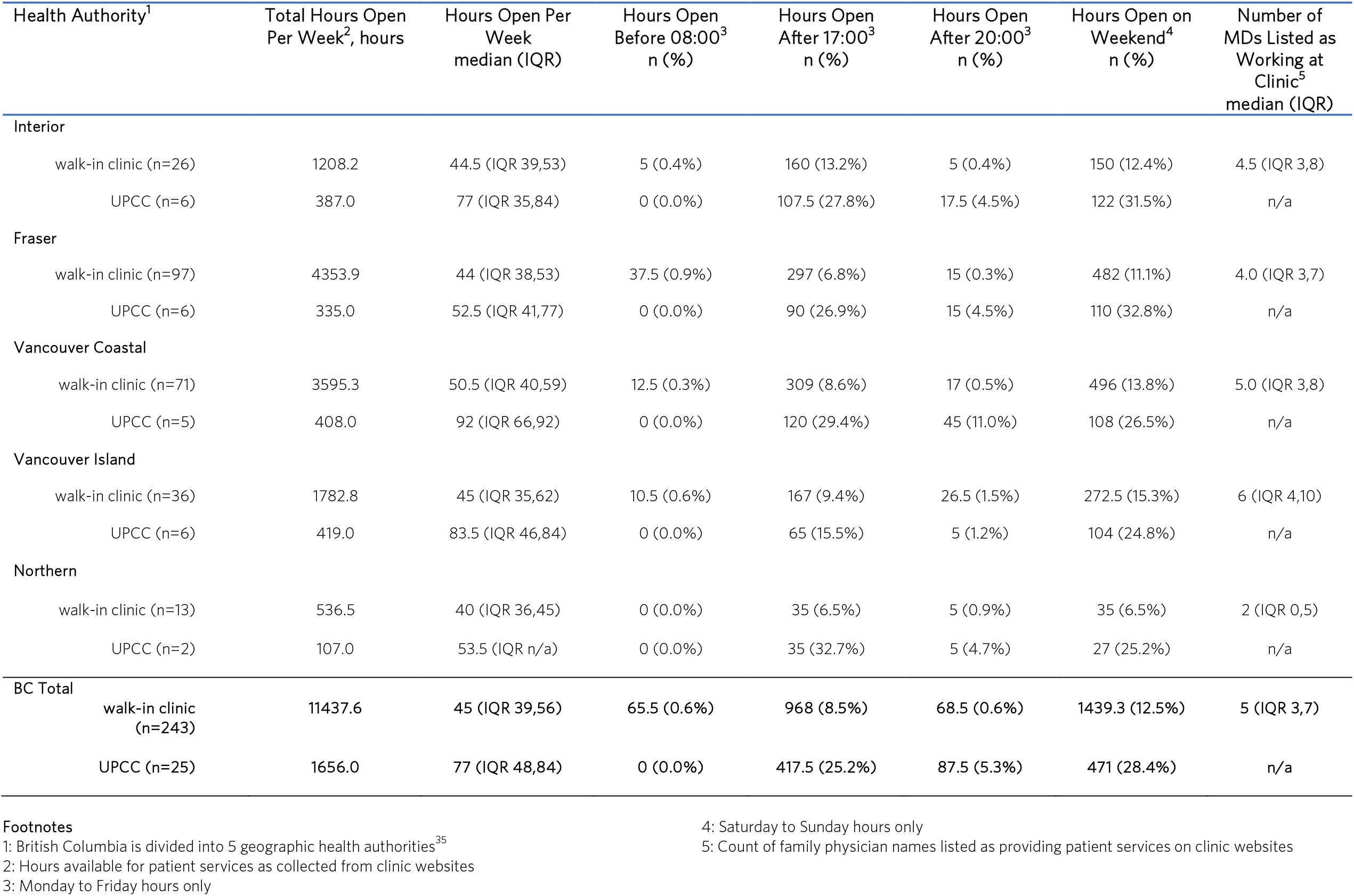
Clinic Service Hours for Walk-in Clinics and Urgent Primary Care Centres in British Columbia

### After hours availability, association with other primary care services, booking methods and appointment types

Table 2 compares primary care access characteristics between walk-in clinics and UPCCs and shows that there are significant differences between the types of clinics, including being associated with longitudinal practice, appointment booking and virtual appointment options. Most walk-in clinics in BC (92.6%) are operated as a part of, or adjacent to, a longitudinal family practice and the majority of hours of service (78.4%) are provided during weekday business hours (08:00-17:00). Further, they offer phone (99.6%) and online booking (46.9%) and virtual appointments (73.3%) at significantly higher rates than UPCCs (44%, 0% and 20% respectively).

**Table 2:** Access Characteristics of Walk-in Clinics and Urgent and Primary Care Centres (UPCCs)

| Access Characteristic | walk-in clinics<br>(n=244)<br>n (%) | UPCCs<br>(n=25)<br>n (%) | X sq | p-value |
| --- | --- | --- | --- | --- |
| Clinic is associated with a longitudinal primary care practice | 225 (92.6%) | 2 (8.0%) | 125.18 | <.001 |
| Patient may walk-in<sup>1</sup> | 223 (91.8%) | 20 (80.0%) | 3.712 | 0.054 |
| Notice of change to walk-in access service due to COVID-19<sup>2</sup> | 108 (44.4%) | 3 (12.0%) | 9.834 | 0.002 |
| **Appointment booking<sup>3</sup>** |  |  |  |  |
| Walk-in only | 1 (0.4%) | 14 (56.0%) | 132.568 | <.001 |
| Phone | 242 (99.6%) | 11 (44.0%) | 143.580 | <.001 |
| Online | 114 (46.9%) | 0 (0.0%) | 20.410 | <.001 |
| Email Bookings | 19 (7.8%) | 0 (0.0%) | 2.104 | 0.147 |
| **Virtual appointments available<sup>4</sup>** | 178 (73.3%) | 5 (20.0%) | 29.680 | <.001 |
| Phone appointments | 176 (72.4%) | 5 (20.0%) | 28.419 | <.001 |
| Video appointments | 136 (56.2%) | 3 (12.0%) | 17.736 | <.001 |
| Pharmacy attached to clinic | 62 (25.5%) | 3 (12.0%) | 2.254 | 0.133 |
| Only open between 08:00-17:00, M-F | 65 (26.7%) | 1 (4.0%) | 6.32 | 0.012 |
| 1: Patient does not need to be previously known, nor have a booked appointment with clinic<br>2: Website included a description of how patient services access has changed due to COVID<br>3: Methods by which a patient can book an appointment with the clinic<br>4: Virtual appointments are available at this location |  |  |  |  |

### Geographic variation in availability of clinic service hours

Figure 1 Describes and compares the total clinic service hours per 100,000 population between walk-in clinics and UPCCs across all 5 geographic health authorities. Interior and Northern Health Authorities cover large areas of less densely populated portions of the province and have lower hours per 100,000, than Fraser, Vancouver Coastal and Vancouver Island Health Authorities, which each contain at least one large metropolitan area. In all regions, the majority of available hours for episodic primary care are provided by walk-in clinics.

**Figure 1:**
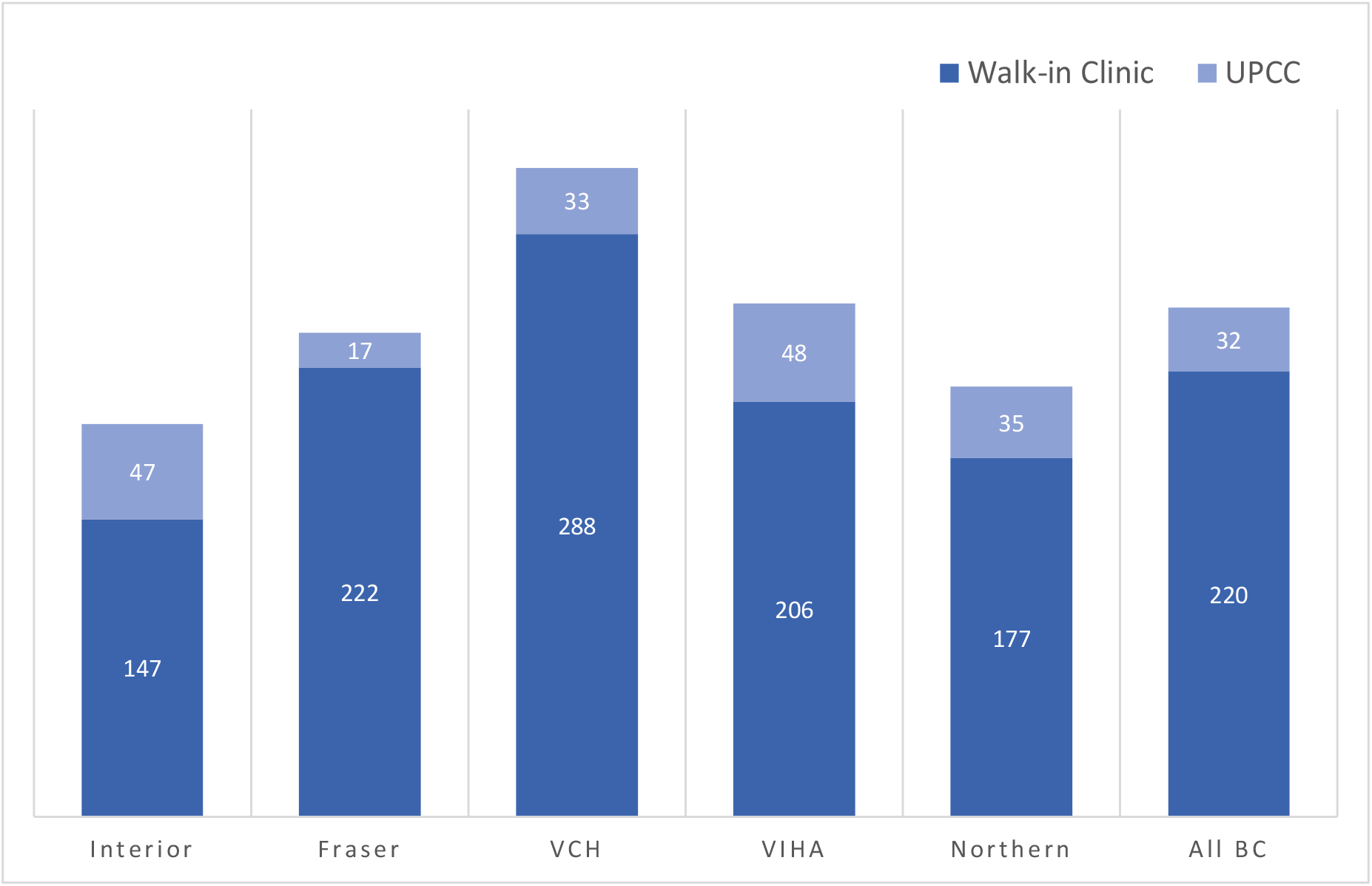
Variation in Number of Clinic Service Hours Available Per Week, Per 100,000 Population, by Health Authority. Footnotes: British Columbia is divided into 5 geographic health authorities: Interior, Fraser, Vancouver Coastal (VCH), Vancouver Island (VIHA), and Northern Population estimates for each health authority were collected from BCStats BC Populations Estimates and Projections^34^

## DISCUSSION

The study was able to describe and compare access characteristics of two types of episodic primary care in British Columbia using publicly available data. The majority of episodic care is provided by walk-in clinics and there is variation between the characteristics present for walk-in clinics versus UPCCs.

### Most walk-in clinics are associated with a longitudinal family practice

Almost all walk-in clinics in BC (92.6%) are operated at the same site as a longitudinal family practice; it is unclear why this structure has emerged as opposed to expansion of longitudinal care given the community need. It is possible resources used to provide episodic care have decreased resources available to provide longitudinal care.^23^ While the original intention of walk-in clinic services may have been to serve as an adjunct to longitudinal care,^24,39^ given the family doctor shortage, it is possible that the current use of walk-in clinics is primarily by people unattached to a longitudinal practice.^22^

In BC, the provincial licensing body, the College of Physicians and Surgeons dictates that walk-in clinics must fulfill the same Practice Standard^7^ as clinics that provide longitudinal care. Notably, this practice standard references continuity but only as it applies to follow up on episodes of medical care and laboratory tests rather than the establishment of a longitudinal relationship between patient and provider that has been shown to reduce mortality.^15^ Across Canada, the practice standards put forward by the provincial and territorial Colleges show variation (See Supplement 1). It is possible that the function and role of walk-in clinics also has variation province to province, and that this clinic type shifts, based on unmet needs of the region it is operating in. Future studies using clinics as a unit of analysis would benefit from transparent reporting of access features to allow for inter-provincial comparison.^40^

### UPCCs offer patient services after-hours more often than walk-in clinics

UPCCs are offering patient services after-hours at higher rates than walk-in clinics. This features is more reminiscent of the original description of walk-in clinic services.^1,2^ The impact of after-hours episodic primary care has had limited study and the authors are unaware of studies examining the impact on the availability of longitudinal care; the authors note that studies looking at costs and emergency department utilization have mixed results.^13,41,42^ Findings from these studies urge careful consideration of local context before creating policies and/or adding resources to increase episodic after-hours access versus longitudinal primary care.^9,11,43^

### Geographic variation

Access to episodic primary care accessibility shows geographic variation in B.C. The province operates using the health authority (HA) system; where each HA is responsible for identifying and meeting their populations health needs.^35^ It is possible that primary care attachment, continuity of care, and after-hours service provision differs according to need across each health authority, or there could be insufficient resources available in some areas. The authors are unaware of any published studies to estimate the “appropriate” amount of episodic primary care for a region. A 2000 study found that rural patient populations were more likely to contact their longitudinal primary care providers during after-hours, compared to those situated in urban areas.^44^

### Limitations

Limitations of this study include the reliance on publicly available data, sourced directly from walk-in clinic and UPCC websites that may not be updated, nor reflect actual practices. Secondly, this study could only measure access to episodic primary care via clinic’s business hours, methods of booking appointments, adjacency to a pharmacy, and availability of virtual services. There was inconsistent reporting of the number of clinicians at work during opening hours and none provided the volume of patients seen per shift. This means that our data cannot be used to indicate the volume of patients seen on a daily basis which would be a more ideal report of service capacity. Despite these limitations, we have been able to use publicly available data to provide a description of episodic primary care access across BC which may be useful to establish a baseline measure in other regions to inform future research, evaluation or policy development of the capacity and impact of episodic primary care.

## Conclusion

There are regional variations in the number of episodic primary care clinic service hours per 100,000 population that may describe underservicing or be reflective of unique local context. Most walk-in clinics are associated with a longitudinal family practice and provide their services during typical business hours. Future studies that include patient care information found in administrative data, and more comprehensive study of access characteristics analyzed at the clinic level would be useful to support the optimization of both episodic and longitudinal primary health care delivery.

## Supporting information

STROBE checklist

## Data Availability

All data produced in the present study are available upon reasonable request to the authors.

## Acknowledgments

Dr. Klimas and Dr. RK McCracken’s work on this paper was supported, in part, by the Canadian Institutes of Health Research.

