## Supplement 1 for "Access to Episodic Primary Care: A Cross-sectional Comparison of Walk-in Clinics and Urgent Primary Care Centers in British Columbia"

### Comparison of College Walk-in Clinic Practice Standards Across Canada

Following initial data collection in British Columbia (BC), policy documents referring to standard of care requirements at walk in clinics Canada were reviewed to determine if there was a consistency in standards of practice and policies governed by each province and territory's College of Physicians and Surgeons or Medical Council. [1-10] We initially searched through each province or territory's College of Physician and Surgeon's or Medical Council's website where we found information from eight provinces and one territory. We then sent an email to the College for each individual province and territory, requesting direction to publicly available information regarding the practice standards for walk-in clinics for primary care within their respective jurisdictions. In our email, we asked each College and Council to identify standards applicable to walk-in clinics that included:

- Clarification to patients about expectations regarding continuity of care.
- Communication with patients regarding abnormal results from investigations ordered by a physician at a walk-in clinic.
- Documentation standards.
- Patient safety and care quality expectations, and;
- The responsibilities for medical administration and/or clinical oversight.

### Results

We received replies from all ten provinces and two of the three territories in Canada. Through personal correspondence with the Charge Physician of Nunavut, it was confirmed that there are no walk-in clinics in this territory and this role is filled by the health centers and the emergency department in Iqaluit. From the policies in place at time of data collection, common practices observed included that the walk-in service providers must: collect a thorough health history and assessment, identify the patient's regular primary care provider, and that they must ensure appropriate follow up and continuity of care.

A detailed comparison of provincial and territorial standards of practice and expectations can be found in Supplementary Table 1.

Supplementary Table 1: Practice Guidelines for Walk-in Clinics and Episodic Care Across Canada Compared to British Columbia

| Province | Practice Guidelines: Has a document or policy for walk-in clinics or episodic care | States that physicians are expected to meet the professional standard of practice | Must collect and document a thorough health history and current health problems | Must identify PCP* and provide summary of episodic interaction | Must inform patient that the regulated member will not provide ongoing care | Patient's who attend clinic repeatedly (and do have a PCP) must be assumed to receive primary health care from that clinic | Clinic has a designated Medical Director | Physicians must ensure appropriate follow up and continuity of care | Must ensure after-hours** coverage is available | Must have on-site access to provincial prescribing records and/or EHR*** for document-tation | Must review medical record and history prior to prescribing controlled substances |
|---|---|---|---|---|---|---|---|---|---|---|---|
| **British Columbia** [3,11] | X | X | X | X | X | X | X | X | X | X | X |
| Alberta[1,12–14] | X | | X | X | X | X | | X | X | X | |
| Saskatchewan [6,15] | X | X | X | X | | X | | X | X | | X |
| Manitoba [2] | | | X | X | | | | X | | | |
| Ontario [4,16] | X | X | X | X | X | | | X | X | | |
| Quebec**** [17,18] | | X | X | | | | | X | | | |
| New Brunswick [8] | X | X | X | X | X | | | X | | X | |
| Newfoundland and Labrador [9] | | X | X | X | | X | X | X | | | |
| Prince Edward Island [5] | X | X | X | X | X | | | X | X | X | X |
| Nova Scotia [7] | X | | X | X | X | | | X | | | X |
| Yukon [10] | X | | X | X | X | | | X | | | X |
| Northwest Territories ***** | | | | | | | | | | | |
| Nunavut ****** | | | | | | | | | | | |

Footnotes

*: PCP: primary care provider
**: After-hours coverage was interpreted as the health care professionals ensuring the patients could go to a different health care facility when their walk-in clinic was closed
***: EHR: electronic health record
****: College de medicines du Quebec does not have specific guidelines for walk-in clinics as per personal correspondence with the Investigations Director of the College
*****: No relevant information could be found from the Northwest Territories Standards of Practice or their Health and Social Services government website.[19] They did not respond to our general inquiry surrounding walk-in clinics and episodic care
******: As per personal correspondence with the Charge Physician of Nunavut, it has been confirmed that there are no walk-in clinics in the territory and this role is fulfilled by the health centers and the emergency department in Iqaluit

17. College des Medicines. Code of ethics of physicians. 2017;20.

18. College des Medicines. Record keeping by physicians in non-hospital settings. 2013 Jun;47.

19. Northwest Territories Health and Social Services Professionals. Community Health Core Service Standards and Protocols [Internet]. Government of the Northwest Territories; 2015 Feb [cited 2021 May 7]. Available from: https://www.hss.gov.nt.ca/professionals/en/services/policies-and-guidelines-standards-and-manuals
